# *Desulfovibrio* Bacteremia in Older Patients with Abdominal Infections, Japan, 2020–2025

**DOI:** 10.1101/2025.09.26.25336699

**Authors:** Naoki Watanabe, Tomohisa Watari, Yoshihito Otsuka

## Abstract

We retrospectively identified eight episodes of *Desulfovibrio* bacteremia at a single tertiary-care hospital in Japan between 2020 and 2025. All patients were ≥65 years of age, and most had comorbidities. The primary source of the infection was the abdomen. The isolates included two *D. desulfuricans*, *D. fairfieldensis*, *D. falkowii*, and *D. legallii*. Matrix-assisted laser desorption/ionization time-of-flight mass spectrometry identified only *D. desulfuricans* and *D. fairfieldensis*, whereas sequence-based identification was required for other species. The minimum inhibitory concentrations (MICs) were low for ampicillin/sulbactam and metronidazole and higher for penicillin, piperacillin/tazobactam, and cefoxitin. Isolates with elevated cephalosporin MICs harbored β-lactamase genes, including *bla*DES-1-like, *bla*CfiA-like, and *bla*MUN-1. Curved or spiral gram-negative rods in anaerobic blood culture bottles, together with a positive desulfoviridin assay, should incite suspicion of *Desulfovibrio* bacteremia of gastrointestinal origin. Sequence-based identification improves the recognition of less common species.

## INTRODUCTION

*Desulfovibrio* are gram-negative, sulfate-reducing, obligately anaerobic, curved or spiral rods that inhabit the soil and aquatic environments as well as the human and animal gut (*1–3*). Human infections are uncommon and have not been completely characterized, with cases manifesting bacteremia (*4–6*) and intra-abdominal infections, such as abscesses and cholecystitis (*7–9*). Less frequent focal infections, include septic arthritis, brain abscess, infected renal cyst, infected arterial aneurysm, and prosthetic joint infection (*10–14*).

The genus *Desulfovibrio* comprises more than 20 species, of which at least six have been implicated in human diseases: *D. desulfuricans*, *D. fairfieldensis*, *D. vulgaris*, *D. piger*, *D. legallii*, and *D. intestinalis* (*15*,*16*). *D. desulfuricans* accounts for the majority of cases with bacteremia, which possibly originates following translocation from the gut (*5*). Although less commonly recognized species, such as *D. legallii*, can cause bloodstream infections, this may be overlooked because matrix-assisted laser desorption/ionization time-of-flight mass spectrometry (MALDI-TOF MS) libraries frequently lack spectra for these species (*17*). Therefore, species-level identification remains uncertain and, even at the genus level, may prove difficult in laboratories.

The optimal therapy for *Desulfovibrio* infections has not yet been established. However, clinical isolates are generally susceptible to metronidazole; carbapenems show some bactericidal activity, whereas they have reduced susceptibility to some β-lactams (*18*). Some *D. desulfuricans* strains produce DES-1, which is an Ambler class A extended-spectrum β-lactamase (*19*). The species-resolved antimicrobial susceptibility data and resistance determinants remain sparse beyond those for *D. desulfuricans* and *D. fairfieldensis.* In this study, we aimed to define the limits of routine identification and provide more consistent recognition and management of underrecognized *Desulfovibrio* species in clinical practice. Thus, we evaluated the performance of MALDI-TOF MS for species-level identification. Moreover, the species were identified using 16S rRNA and whole-genome sequencing. We summarized the antimicrobial susceptibility, resistance determinants, and patient characteristics and outcomes.

## Materials and Methods

### Study design and data collection

This retrospective cohort study was conducted at a single tertiary care hospital in Japan for cases treated between January 2020 and June 2025. We identified cases by searching the laboratory information system for blood cultures that were positive for *Desulfovibrio* spp. All positive sample bottles collected during the same clinical episode were attributed to a single case. We assigned a new episode only when it was evidently based on new symptoms or signs, a new anatomic focus, or following the resolution of a prior episode.

Clinical and laboratory data were extracted from the electronic medical records and laboratory systems. The clinicodemographic variables included age, sex, acquisition type, comorbidities, presenting symptoms, presumed source, antimicrobial therapy, and discharge outcomes. Microbiological variables included the number of blood culture sets that were obtained, number of positive sets, time to positivity, and identification results. We defined the time to positivity as the interval from the start of incubation to the first instrument-flagged positive bottle. We defined healthcare-associated infection as onset ≥48 h after admission or infection that occurred during ongoing medical care, including nursing-home residence, dialysis, or outpatient chemotherapy. All other infections were categorized as community-acquired infections. To contextualize our findings, we reviewed the published cases of *Desulfovibrio* bacteremia. We searched PubMed using the query “*Desulfovibrio* AND bacteremia.” We included English-language case reports, case series, and research articles in which *Desulfovibrio* spp. Were isolated from blood cultures. The patient characteristics and microbiological data were extracted using the same variable definitions as those that we used for our cohort. This study was approved by the Ethics Committee of Kameda Medical Center (Approval No. 25-061). The requirement for informed consent was waived owing to the retrospective design and use of deidentified data.

### Blood culture and identification testing

Blood cultures were processed using the BD BACTEC FX system (BD Biosciences, https://www.bd.com). BACTEC Plus Aerobic/F and Lytic/10 Anaerobic/F bottles were used for adults, whereas BACTEC Peds Plus/F bottles (BD) were used for pediatric patients. The bottles were incubated at 35°C for up to 7 days. *Desulfovibrio* isolates were stored at −80°C. Before testing, the isolates were subcultured on Brucella HK agar (Kyokuto Pharmaceutical Industrial, Tokyo, Japan; https://www.kyokutoseiyaku.co.jp) and incubated anaerobically at 35–37°C for 4–6 days with the AnaeroPack system (Mitsubishi Gas Chemical Company, https://www.mgc.co.jp).

Species identification by MALDI-TOF MS was performed using a MALDI Biotyper with the MBT Compass Library version 13 (Bruker Daltonics GmbH, https://www.bruker.com). We accepted species-level identifications at scores ≥2.0 whereas a score <2.0 was defined as an uncertain MALDI result. Catalase and desulfoviridine assays were performed for biochemical testing (Appendix Methods).

### 16S rRNA gene sequencing

We used 16S rRNA gene sequencing and whole-genome analyses to identify the species. Genomic DNA was extracted using a MagLEAD instrument along with the manufacturer’s kit (Precision System Science, https://www.pss.co.jp), with primers that were previously reported (*20*,*21*). The primer sequences and cycling conditions are provided in Appendix Table 1. The sequences were trimmed and queried using BLASTn, restricting the search set to the material type. For 16S-based species assignment, percent identity ≥99.0% was considered a supportive finding (*22*).

**TABLE 1.** Clinical characteristics of patients with *Desulfovibrio* bacteremia in a single tertiary-care hospital, Japan, 2020–2025.

| Characteristics | All (n = 8) |
| --- | --- |
| Age, median y (IQR) | 81 (77, 86) |
| Sex |  |
| Female | 2 (25) |
| Male | 6 (75) |
| Charlson Comorbidity Index |  |
| Score 0–1 | 1 (13) |
| Score 2–4 | 6 (75) |
| Score $\geq 5$ | 1 (13) |
| Comorbid conditions |  |
| Solid tumor | 3 (38) |
| Hematologic malignancy | 1 (13) |
| Immunocompromised condition | 1 (13) |
| Recent antimicrobial use ($\leq 30$ days) | 5 (63) |
| Acquisition type |  |
| Healthcare-associated | 4 (50) |
| Community-acquired | 4 (50) |
| Source of infection |  |
| Abdominal | 7 (88) |
| Unknown | 1 (13) |
| ICU admission | 1 (13) |
| Septic shock | 1 (13) |
| In-hospital death | 1 (13) |

**Footnote:** Data are no. (%) unless otherwise indicated. ICU, intensive care unit. IQR, interquartile range.

Multiple sequence alignment was performed using MUSCLE in MEGA version 12 (*23*). We constructed a neighbor-joining tree using the Kimura 2-parameter model, pairwise deletion of gaps, and 100 bootstrap replicates. Using a prespecified ≥1,300-bp length threshold, we reanalyzed the 16S sequences that were previously reported as *D. fairfieldensis*. Two sequences met this criterion (strain FH 26001/95, U42221.1; strain D4, AF192155.1) and were analyzed alongside the isolates from this study. Trees were visualized and annotated using Interactive Tree of Life (iTOL) version 7 (*24*).

### Whole-genome sequencing and analysis

Libraries were prepared using Illumina DNA Prep (M) Tagmentation (Illumina, https://www.illumina.com), and a MiSeq instrument was used to generate paired-end reads. These reads were assembled de novo on a Bacterial and Viral Bioinformatics Resource Center platform (*25*). The assembly quality was evaluated using CheckM (*26*). Genomes were annotated using DFAST (*27*). Species assignment was resolved by average nucleotide identity (ANI) that was computed using fastANI (*28*) against type strains and the reference genomes from the Genome Taxonomy Database. For *D. fairfieldensis*, we performed digital DNA–DNA hybridization using a Genome-to-Genome Distance Calculator (*29*). Antimicrobial resistance determinants were identified from the draft assemblies by using AMRFinderPlus (*30*). Details of the tools used for whole-genome analysis are provided in Appendix Table 2.

**TABLE 2.**
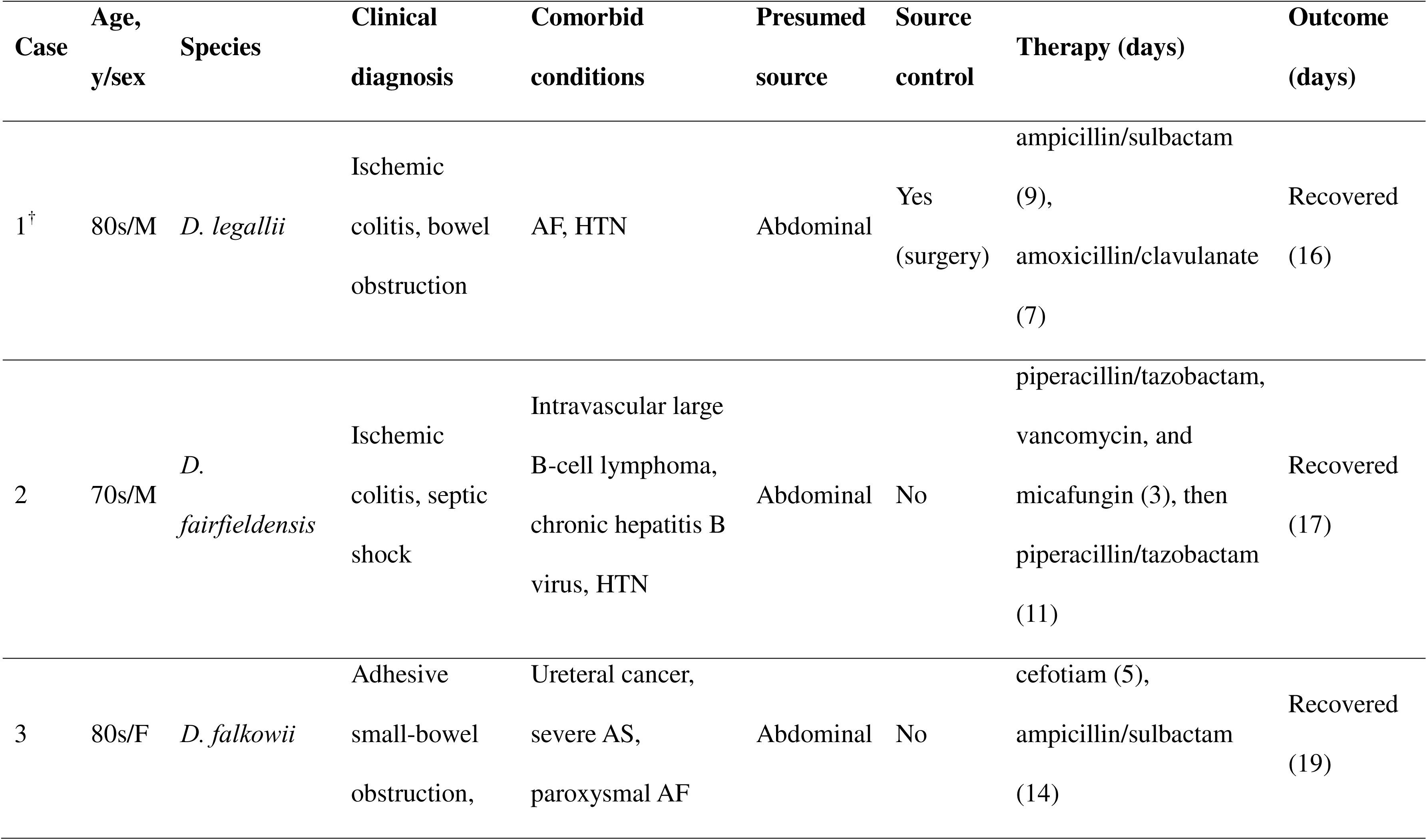
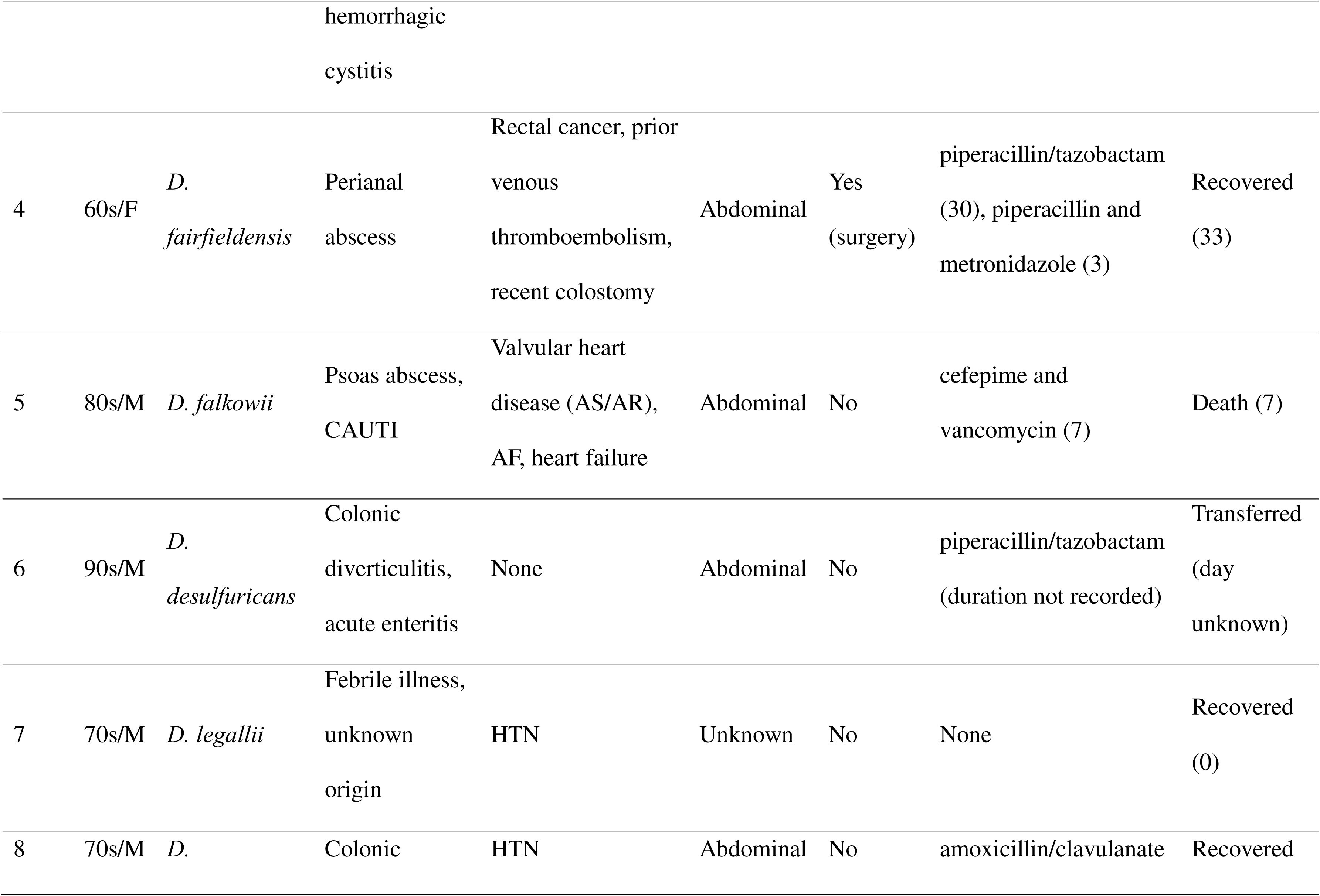
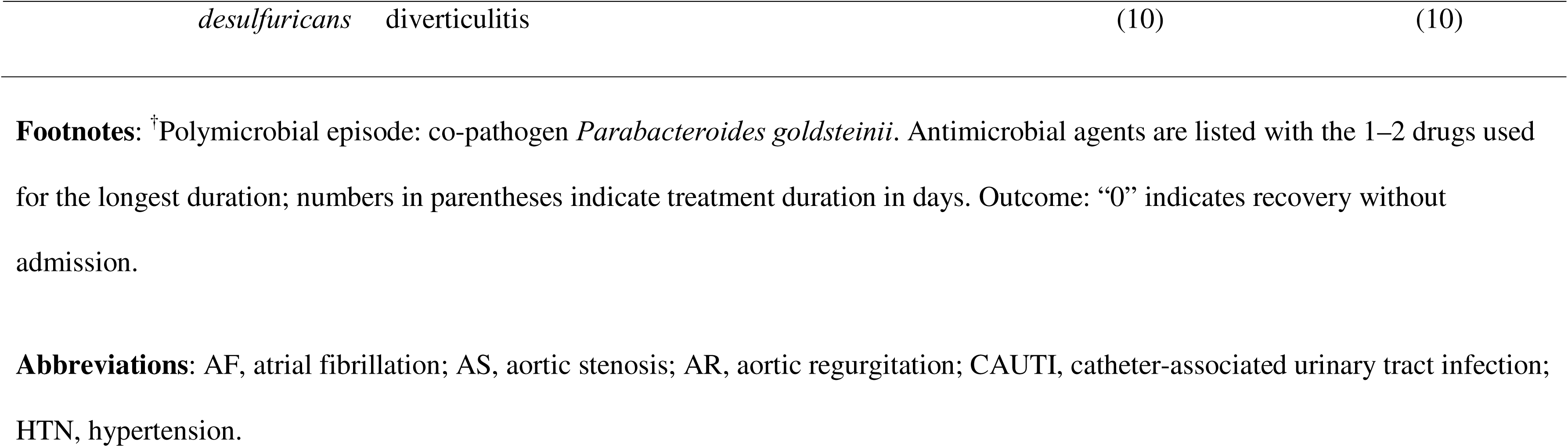
Patient characteristics and clinical course of *Desulfovibrio* bacteremia in a single tertiary-care hospital, Japan, 2020–2025.

### Antimicrobial susceptibility testing

Antimicrobial susceptibility was determined by microdilution in Brucella broth on dry plates (Eiken Chemical, https://www.eiken.co.jp). The inocula were prepared according to the manufacturer’s instructions and dispensed at a final concentration of 1×10^5^ CFU per well. The plates were incubated anaerobically at 35–37°C for 46–48 h. In case of insufficient growth in the control well, the incubation was extended for up to 96 h. After confirming adequate growth in the controls, the minimum inhibitory concentrations (MIC) were read as the lowest concentrations without visible growth. Susceptibility to penicillin, ampicillin/sulbactam, amoxicillin/clavulanate, piperacillin/tazobactam, ceftriaxone, cefoxitin, imipenem, clindamycin, moxifloxacin, and metronidazole was tested. As the CLSI/EUCAST breakpoints have not been established for *Desulfovibrio* by broth microdilution, we reported the MICs in only μg/mL.

### Statistical analysis

The analyses were performed using EZR version 1.54 (*31*). Continuous variables were summarized as medians with interquartile ranges and, when appropriate, as ranges. Categorical variables were summarized as counts and percentages and reported as n/N (%). Two-sided 95% confidence intervals (CIs) for proportions were calculated using the exact Clopper–Pearson method. Given the small sample size, all analyses were descriptive and no formal hypothesis testing was planned.

## RESULTS

### Patient characteristics and clinical courses

During January 2020–June 2025, we confirmed 8 episodes of *Desulfovibrio* bacteremia, which were detected in 8 out of 4,431 patients with positive blood cultures (0.2% [95% CI 0.1%–0.4%]). Table 1 summarizes the patient characteristics. Appendix Table 3 provides detailed information on the clinical variables and comorbidity data. No episodes of recurrence were observed. The median age was 81 (IQR 77–86) years. All patients were at least 65 years old. Six patients were male and two were female. Seven patients had a Charlson Comorbidity Index score ≥2. The presumed source of infection was the abdomen in seven cases and was unknown in one case.

**TABLE 3.**
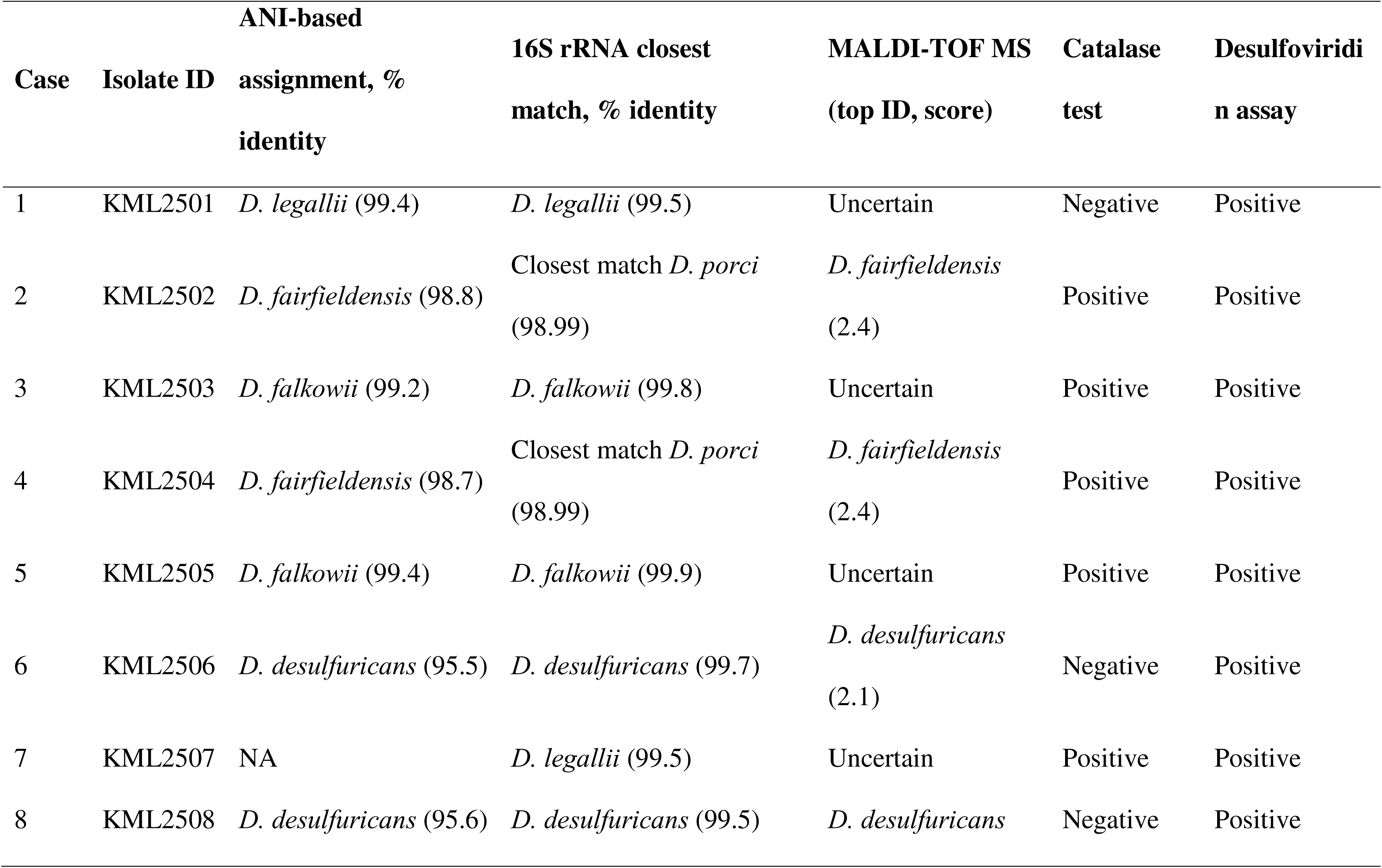
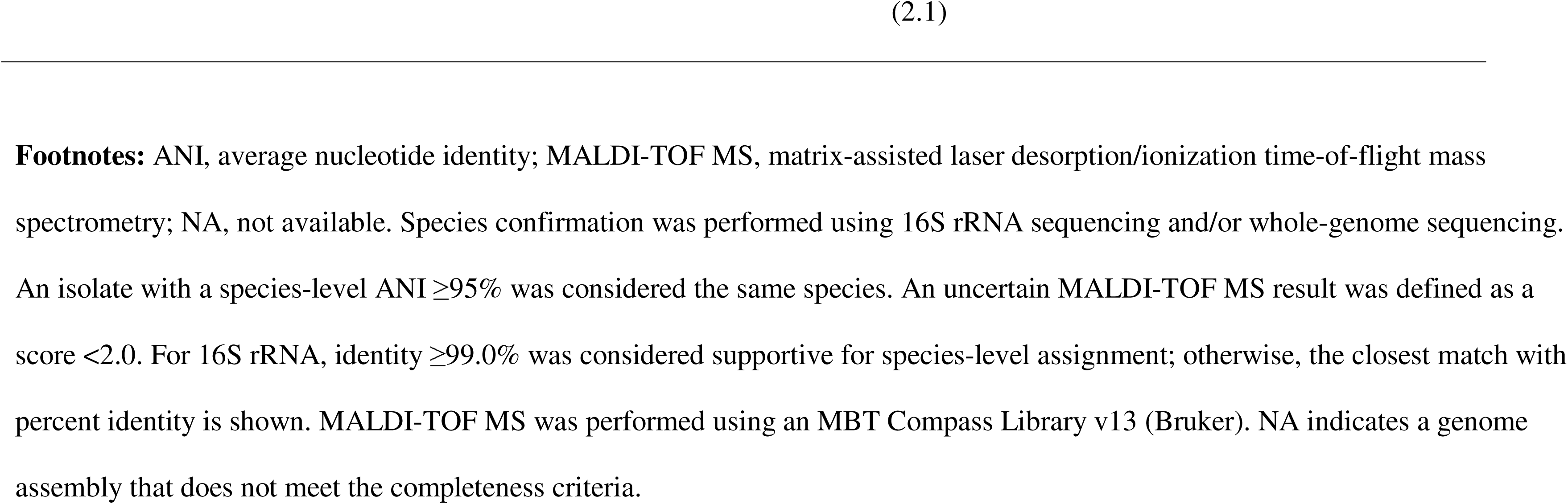
Identification of *Desulfovibrio* in isolates from patients with bacteremia in a single tertiary-care hospital, Japan, 2020–2025.

The most common presenting symptoms were fever (4/8 [50%]) and abdominal symptoms (5/8 [63%]). The median time to positivity was 4 (range: 2–6) days. In three cases, the bottles tested positive on day 5 or later. One episode was polymicrobial, with *Parabacteroides goldsteinii* identified as a co-pathogen.

### Clinical outcomes

Seven of the eight patients were diagnosed with true bacteremia and received antibiotics. The most frequently used agents were β-lactam/β-lactamase inhibitor combinations as follows: piperacillin/tazobactam in 3 patients, ampicillin/sulbactam in 2, and amoxicillin/clavulanate in 2. One patient (Case 7) did not receive antibiotics because he was asymptomatic at the time of culture positivity and declined further evaluation.

The patterns of *D. falkowii* and *D. legallii* were similar to those of *D. desulfuricans* and *D. fairfieldensis*, with infection in older adults that predominantly originated from abdominal sources. *D. falkowii* bacteremia occurred in two patients in their 80s, and both were presumed to have originated from abdominal sources (cases 3 and 5). Case 3 had an adhesive small-bowel obstruction and recovered after cefotiam treatment for 5 days, followed by ampicillin/sulbactam treatment for 14 days. Case 5 had a persistent psoas abscess that was not drained; despite cefepime and vancomycin treatment, the patient died on hospital day 7. *D. legallii* bacteremia occurred in two men (cases 1 and 7). Case 1 involved ischemic colitis with a descending colonic stricture and recovered after treatment with ampicillin/sulbactam for 9 days, followed by amoxicillin/clavulanate for 7 days. Case 7 presented with febrile illness of unknown origin. *Desulfovibrio* grew in the blood culture after 6 days of incubation. However, by the time of notification, his symptoms had resolved without antibiotics and he declined further evaluation. The patient was alive at the 1-year follow-up for an unrelated illness.

### Microbiological characteristics and identification testing

Gram staining of positive blood culture bottles revealed gram-negative rods with a curved or spiral morphology (Figure 1). Species assignment by 16S rRNA sequencing and ANI identified two isolates each of *D. desulfuricans*, *D. fairfieldensis*, *D. falkowii*, and *D. legallii* (Table 4 and Figure 2). MALDI-TOF MS yielded of 4/8 correct species-level identifications and 4/8 uncertain results. Correct calls were made for *D. desulfuricans* and *D. fairfieldensis* whereas *D. falkowii* and *D. legallii* produced uncertain results. Catalase was positive in 5 out of 8 isolates (2/2 *D. falkowii*, 1/2 *D. legallii*, and 2/2 *D. fairfieldensis*). The desulfoviridin assay was positive for all isolates.

**Figure 1.**
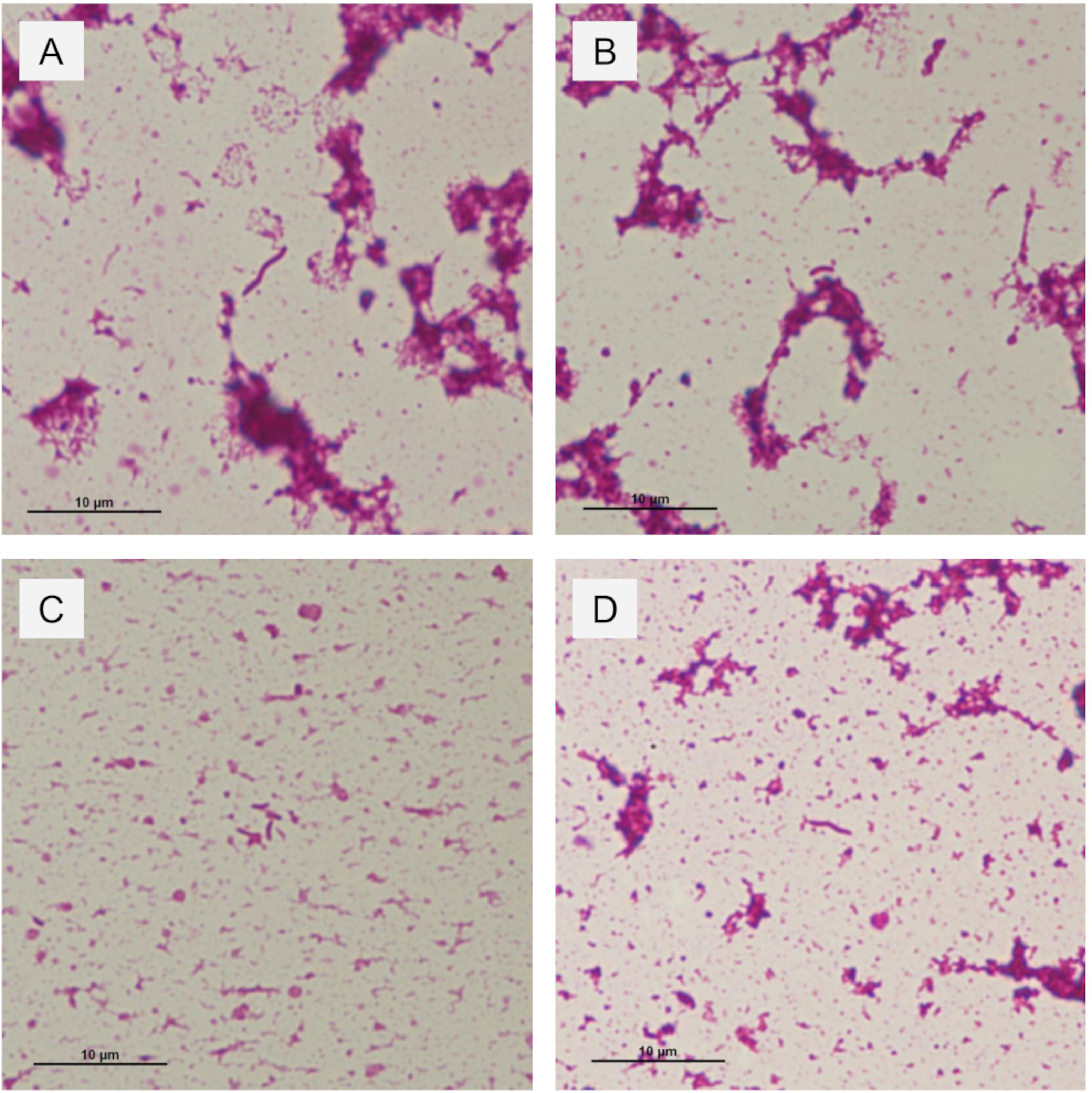
Gram-stained smear from a positive anaerobic blood-culture bottle in a study of *Desulfovibrio* bacteremia at a tertiary-care hospital in Japan, 2020–2025. Curved or spiral gram-negative rods are visible. Panels: A, *D. desulfuricans* (spiral form); B, *D. desulfuricans* (curved form); C, *D. falkowii* (curved form); D, *D. legallii* (spiral form). Images were acquired at 1,000× total magnification using a 100× oil-immersion objective. Scale bar, 10 μm.

**Figure 2.**
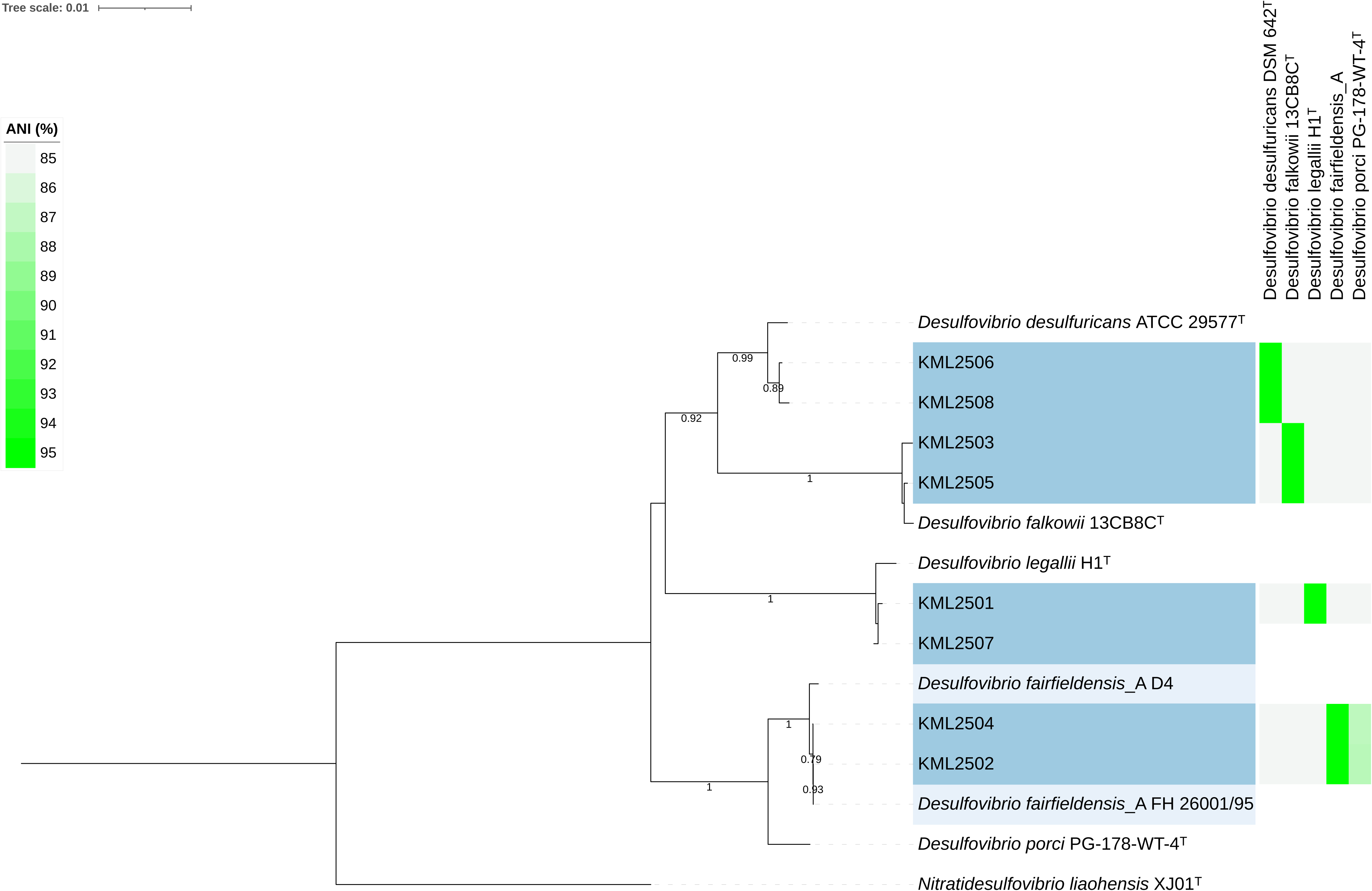
Phylogenetic tree of study isolates within the genus *Desulfovibrio* based on the identification of 16S rRNA gene sequences. Study isolates are marked in blue. Previously reported *D. fairfieldensis* isolates (FH 26001/95 and D4) are marked in light blue. Type strains are marked with a superscript T. The scale bar indicates the number of substitutions per site. ANI heatmap and dendrogram comparing study isolates with type or reference genomes. Cells are colored by ANI (%) on a fluorescent-green gradient, with higher identity appearing brighter. **Abbreviations.** ANI, average nucleotide identity.

**TABLE 4.**
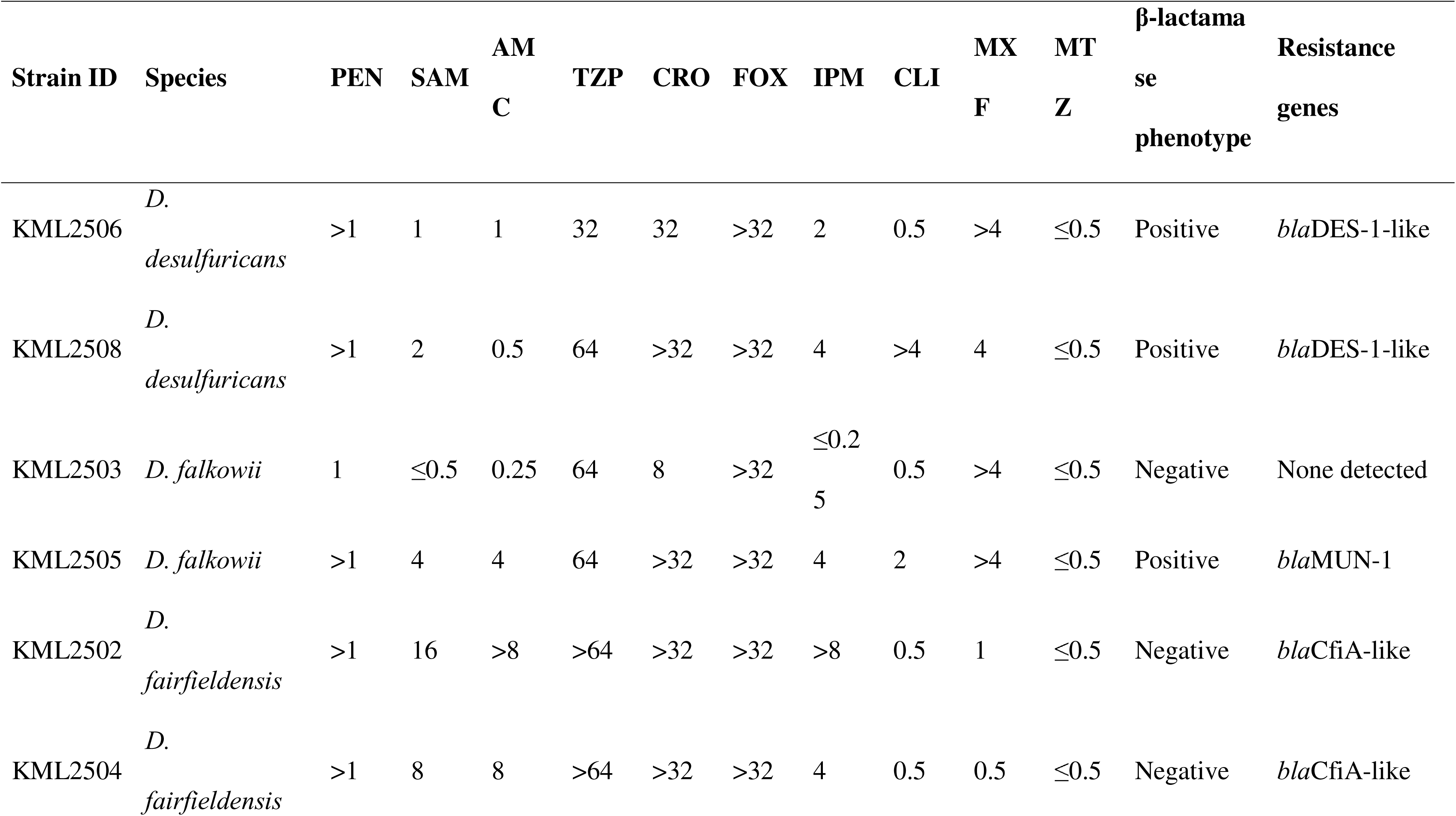
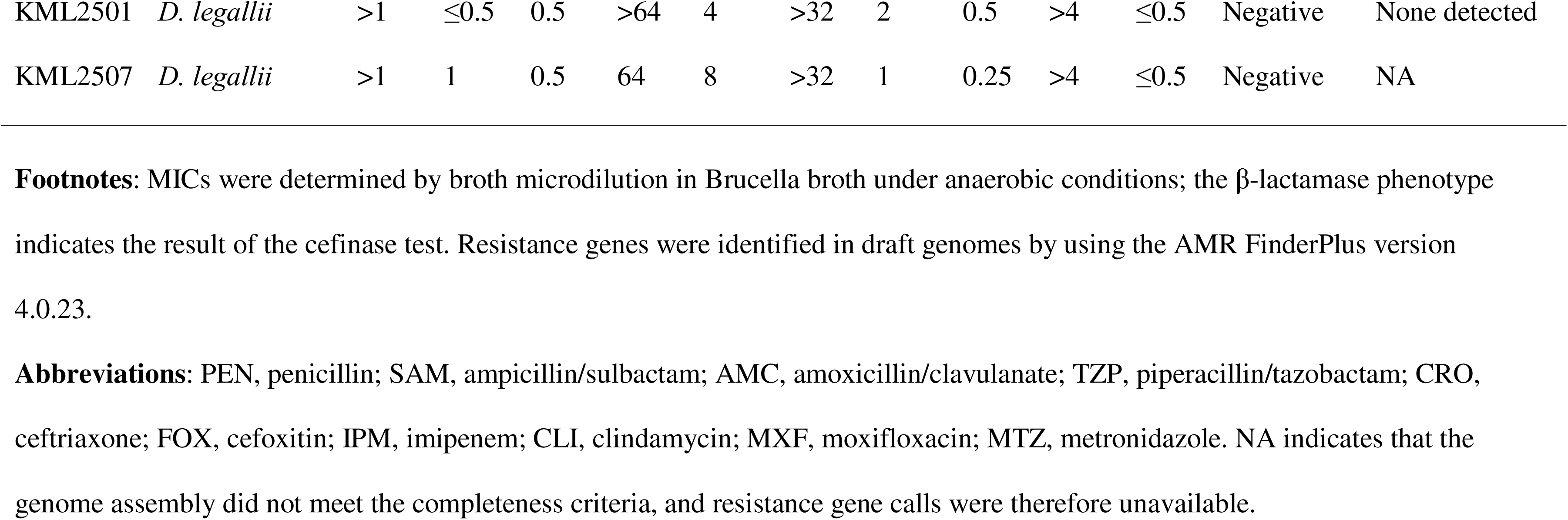
MIC (μg/mL) and resistance genes in *Desulfovibrio* isolates from patients with bacteremia in a single tertiary-care hospital, Japan, 2020–2025.

| Strain ID | Species | PEN | SAM | AM<br>C | TZP | CRO | FOX | IPM | CLI | MX<br>F | MT<br>Z | β-lactamase<br>phenotype | Resistance<br>genes |
| --- | --- | --- | --- | --- | --- | --- | --- | --- | --- | --- | --- | --- | --- |
| KML2506 | *D. desulfuricans* | >1 | 1 | 1 | 32 | 32 | >32 | 2 | 0.5 | >4 | ≤0.5 | Positive | *bla*DES-1-like |
| KML2508 | *D. desulfuricans* | >1 | 2 | 0.5 | 64 | >32 | >32 | 4 | >4 | 4 | ≤0.5 | Positive | *bla*DES-1-like |
| KML2503 | *D. falkowii* | 1 | ≤0.5 | 0.25 | 64 | 8 | >32 | ≤0.2<br>5 | 0.5 | >4 | ≤0.5 | Negative | None detected |
| KML2505 | *D. falkowii* | >1 | 4 | 4 | 64 | >32 | >32 | 4 | 2 | >4 | ≤0.5 | Positive | *bla*MUN-1 |
| KML2502 | *D. fairfieldensis* | >1 | 16 | >8 | >64 | >32 | >32 | >8 | 0.5 | 1 | ≤0.5 | Negative | *bla*CfiA-like |
| KML2504 | *D. fairfieldensis* | >1 | 8 | 8 | >64 | >32 | >32 | 4 | 0.5 | 0.5 | ≤0.5 | Negative | *bla*CfiA-like |
| KML2501 | *D. legallii* | >1 | ≤0.5 | 0.5 | >64 | 4 | >32 | 2 | 0.5 | >4 | ≤0.5 | Negative | None detected |
| KML2507 | *D. legallii* | >1 | 1 | 0.5 | 64 | 8 | >32 | 1 | 0.25 | >4 | ≤0.5 | Negative | NA |

**Footnotes:** MICs were determined by broth microdilution in Brucella broth under anaerobic conditions; the β-lactamase phenotype indicates the result of the cefinase test. Resistance genes were identified in draft genomes by using the AMR FinderPlus version 4.0.23.
**Abbreviations:** PEN, penicillin; SAM, ampicillin/sulbactam; AMC, amoxicillin/clavulanate; TZP, piperacillin/tazobactam; CRO, ceftriaxone; FOX, cefoxitin; IPM, imipenem; CLI, clindamycin; MXF, moxifloxacin; MTZ, metronidazole. NA indicates that the genome assembly did not meet the completeness criteria, and resistance gene calls were therefore unavailable.

Seven genomes met the inclusion criteria of completeness and contamination (Appendix Table 4). One genome was incomplete (<90% completeness) and therefore excluded from subsequent analyses. The genome sizes ranged from 2.8 to 3.5 Mb. ANI resolved seven genomes: *D. desulfuricans* (n=2), *D. fairfieldensis* (n=2), *D. falkowii* (n=2), and *D. legallii* (n=1). Using 16S rRNA gene analysis, the excluded genome was identified as *D. legallii*. For *D. desulfuricans*, *D. falkowii*, and *D. legallii*, 16S assignments were concordant with ANI (95.5%–99.4% for type or reference strains, Appendix Table 5).

**TABLE 5.**
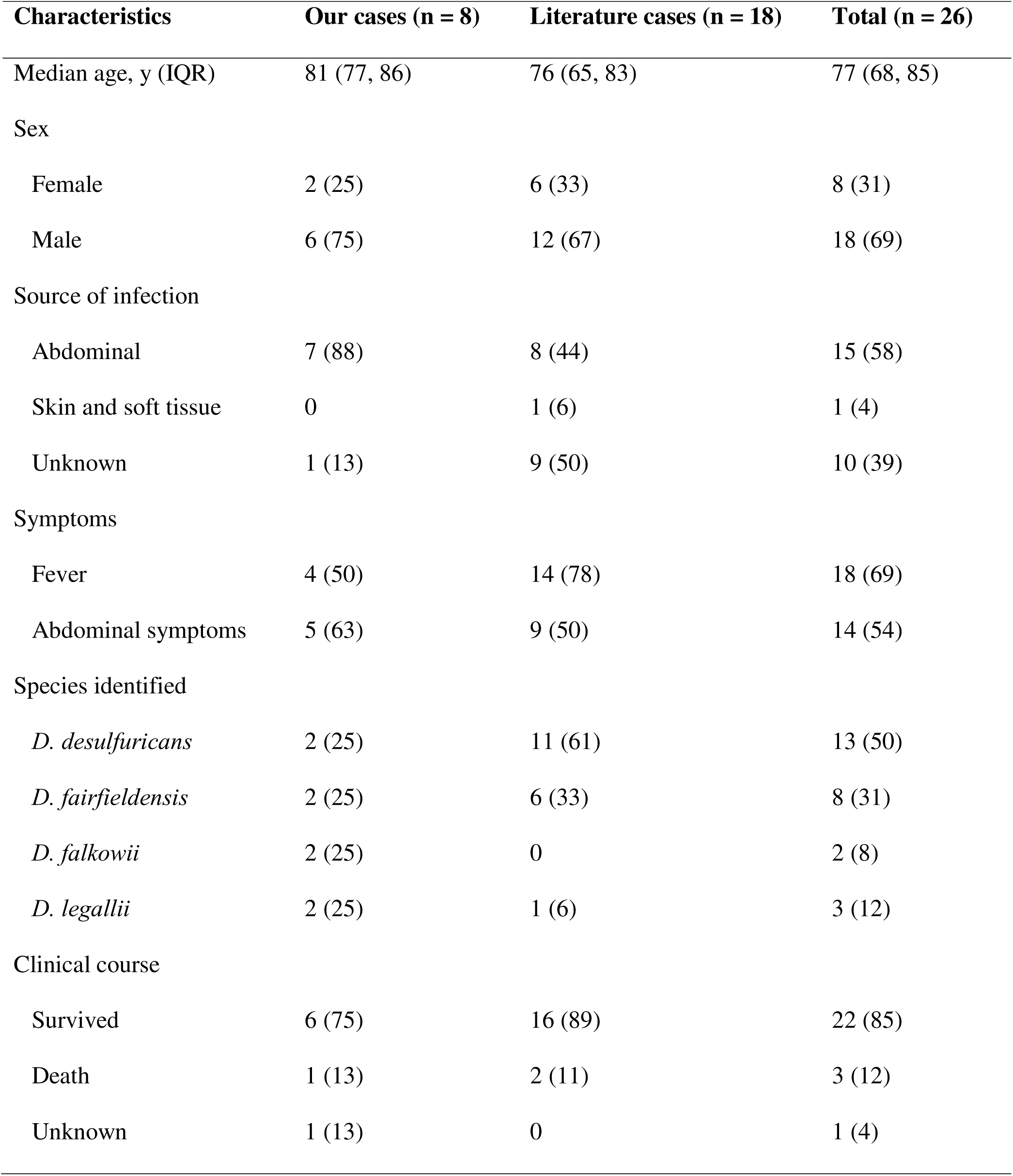
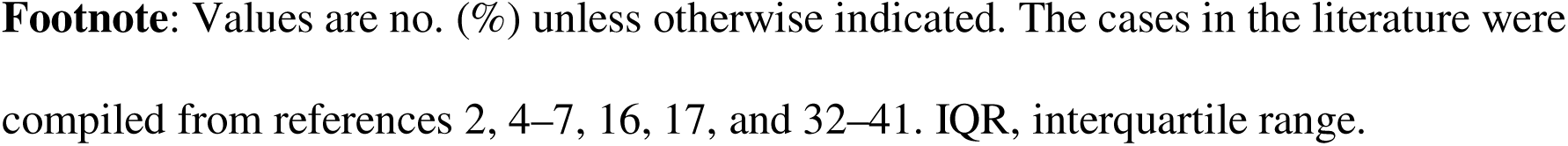
Summary of *Desulfovibrio* bacteremia cases from the study cohort and the literature.

Two isolates (KML2502 and KML2504) were assigned as *D. fairfieldensis* by ANI yet showed 98.99% 16S identity to *D. porci* PG-178-WT-4^T^. dDDH supported the ANI-based assignment: KML2502 and KML2504 showed dDDH values >80% for *D. fairfieldensis* but only 32% for *D. porci*, with a corresponding ANI of 86% for *D. porci*. Similarly, reanalysis of historical *D. fairfieldensis* sequences (strains FH 26001/95 and D4) showed the highest 16S similarity to *D. porci* (98.99%), which was consistent with our *D. fairfieldensis* sequences.

### Antimicrobial susceptibility and resistance genes

Antimicrobial susceptibility results were available for all the eight isolates (Table 4). The MICs were low for ampicillin/sulbactam and metronidazole and high for penicillin, piperacillin/tazobactam, and cefoxitin. Higher MICs were common for ceftriaxone and moxifloxacin, whereas lower MICs were common for amoxicillin/clavulanate, imipenem, and clindamycin. For ceftriaxone, the MICs were 4 to ≥32 μg/mL. Isolates with ceftriaxone MICs ≥32 μg/mL included two *D. desulfuricans* (KML2506 and KML2508), two *D. fairfieldensis* (KML2502 and KML2504), and one *D. falkowii* (KML2505). Phenotypically, the cefinase test was positive for *D. desulfuricans* KML2506 and KML2508 and for *D. falkowii* KML2505.

Genome screening with AMRFinderPlus identified β-lactamase genes in five isolates We did not detect non–β-lactam resistance determinants. *D. falkowii* KML2505 carried *bla*MUN-1 with 100% identity and 100% coverage (reference WP_206340447.1). *D. desulfuricans* KML2506 and KML2508 each carried DES-family class A β-lactamases (*bla*DES-1-like) with 81–82% identity and 100% coverage (closest reference WP_063860095.1). In addition, *D. fairfieldensis* KML2502 and KML2504 harbored subclass B1 metallo-β-lactamase homologs (*bla*CfiA-like) with 47% identity and 94% coverage (closest reference WP_005808062.1). However, under these criteria, no β-lactamase was detected in KML2501 or KML2503.

### Comparison with cases reported in the literature

Including 18 previously published cases (*2*,*4–7*,*16*,*17*,*32–41*), we summarized 26 episodes of *Desulfovibrio* bacteremia (Table 5). The median patient age was 77 (IQR 68–85) years. Males comprised 18 out of the 26 (69%) patients. Abdominal sources were identified in 15 of the 26 (58) patients, whereas 10 of the 26 (39%) patients had no clear source. The most common symptoms were fever in 18 of the 26 (69%) and abdominal symptoms in 14 of the 26 (54%) patients. *D. desulfuricans* was detected in 13 of the 26 (50%) patients, followed by *D. fairfieldensis* in 8 of the 26 (31%) patients. *D. legallii* represented 3 out of 26 (12%) cases, including one previously reported case whereas *D. falkowii* represented 2 of the 26 (8%) cases, which was observed only in our series. The overall mortality rate was 3 out of 26 (12%; 95% CI 2%–30%) and was similar between our cohort 1/8 (13%) and prior reports 2/18 (11%).

## DISCUSSION

Here, we highlight three findings of *Desulfovibrio* bacteremia. Four *Desulfovibrio* species caused bloodstream infection in this cohort. Most episodes appeared to originate in the abdomen. MICs were low for ampicillin/sulbactam and metronidazole and higher for several β-lactams. In this cohort, the species diversity was broader than that in earlier reports, which emphasized *D. desulfuricans* and *D. fairfieldensis* were the predominant causes of bacteremia (*5*). In our study, these two species accounted for half of the cases, whereas *D. falkowii* and *D. legallii* accounted for the remainder. This difference likely reflected the limitations of routine identification. The MALDI-TOF MS library used in our laboratory did not include *D. falkowii* or *D. legallii*, and these species were not resolved at the species level by MALDI-TOF MS. Furthermore, a 2025 report on *D. legallii* bacteremia noted that MALDI-TOF MS failed to identify the species (*17*). Therefore, incorporating the reference spectra of these species into clinical libraries would enhance their identification.

Our data indicated that the 16S rRNA gene sequencing alone does not reliably distinguish *D. fairfieldensis* from *D. porci*. Isolates assigned to *D. fairfieldensis* and previously reported strains showed 98.99% 16S identity with the *D. porci* type strain, and this constitutes a range in which closely related species frequently remain unresolved by 16S analysis (*22*). In contrast, ANI separated *D. fairfieldensis* from *D. porci* in our study, and dDDH supported these assignments, which is consistent with accepted thresholds for ANI ≥95% and dDDH ≥70% (*29*,*42*). From a nomenclatural perspective, *D. fairfieldensis* has not been validly published under the International Code of Nomenclature of Prokaryotes and lacks a type strain (*43*), whereas *D. porci* has been validly described as a type strain (*44*). A practical approach involves the use of updated MALDI-TOF MS libraries and to refer inconclusive isolates for whole-genome analysis. All the patients were older adults. Fever and abdominal symptoms were common, and an abdominal source was presumed in most cases, which supports the gastrointestinal origin of this genus. The clinical features of our cases were consistent with those that were previously reported. Although *D. falkowii* and *D. legallii* are rarely reported as the cause of bacteremia (*17*,*45*), their clinical profiles followed the same pattern as those of *D. desulfuricans* and *D. fairfieldensis* regarding the age, source, and outcome. In routine practice, curved or spiral gram-negative rods in anaerobic blood culture bottles suggest *Desulfovibrio*. The anaerobic-dependent growth and desulfoviridin assays supported this hypothesis. The desulfoviridin assay serves as a genus-level screen for *Desulfovibrio* (*15*). However, as *Bilophila wadsworthia* can also test positive on this screen, the assay is supportive rather than specific. Therefore, the colony morphology and MALDI-TOF MS results should be interpreted.

These antimicrobial susceptibility patterns are consistent with previous observations (*18*,*46*). The MICs were higher for penicillin, piperacillin/tazobactam, and cefoxitin. Isolates with ceftriaxone MICs ≥32 μg/mL carried β-lactamase genes, including *bla*DES-1-like, *bla*MUN-1, and *bla*CfiA-like. These findings suggest that β-lactamase activity contributes to the elevated ceftriaxone MICs in some isolates. *bla*MUN-1 encodes an Ambler class A ESBL that confers cephalosporin resistance (*47*), which is consistent with our observations. *bla*DES-1 encodes an Ambler class A ESBL that was originally described in *D. desulfuricans* (*19*); furthermore, the *bla*DES-1-like assignment is compatible with this mechanism, although the lower identity warrants functional confirmation. For *bla*CfiA-like homologs, the negative cefinase results suggested limited expression under the test conditions. Additional mechanisms, including reduced outer membrane permeability and increased efflux pump activity, may contribute to this outcome.

This study had some limitations. The single-center retrospective design and the small sample size limit the generalizability of the results. One *D. legallii* genome had insufficient completeness and was excluded from the whole-genome analyses. The 16S sequencing assigned the isolate to *D. legallii*, and the 16S assignments for the remaining isolates aligned with those of ANI, and thereby made misclassification unlikely. Functional validation of *bla*DES-1-like and *bla*CfiA-like genes was not performed, and their causal links to MICs remain unconfirmed. Future work should assess β-lactamase inhibitor synergy, gene expression, and enzymatic activity to clarify genotype–phenotype links in *Desulfovibrio*.

In conclusion, *Desulfovibrio* bacteremia involves *D. desulfuricans*, *D. fairfieldensis*, *D. falkowii*, and *D. legallii*. These cases occurred in older adults with presumed abdominal sources. MICs were low for metronidazole and ampicillin/sulbactam and higher for several β-lactams. Routine MALDI-TOF MS did not identify *D. falkowii* or *D. legallii*. Nonetheless, the morphology in anaerobic bottles, together with a positive desulfoviridin assay, can generate suspicion of *Desulfovibrio* infection and guide early management while awaiting confirmatory identification.

## Data Availability

All data produced in the present study are available upon reasonable request to the authors.

## Acknowledgments

We thank Wataru Hayashi for advice on whole-genome analysis. This work received no specific grant from any funding agency in the public, commercial, or nonprofit sectors.

