## Appendix for "*Desulfovibrio* Bacteremia in Older Patients with Abdominal Infections, Japan, 2020–2025"

Appendix Methods

Appendix Table 1. Primers for 16S rRNA gene amplification and sequencing

Appendix Table 2. Details of the tools used for whole-genome analysis and a list of Type strains

Appendix Table 3. Detailed clinical variables and comorbidity data

Appendix Table 4. Genome statistics of *Desulfovibrio* isolates

Appendix Table 5. Sequence-based analysis results

### Appendix Methods

#### Desulfoviridin assay

Apply 2 N NaOH to the swabbed colony or broth sediment and examine immediately under 365-nm UV in a dark box; red fluorescence indicates a positive result.

#### Catalase test

Expose fresh colonies to 15% hydrogen peroxide; immediate effervescence is positive.

### Appendix Table 1. Primers for 16S rRNA gene amplification and sequencing

#### A. Primers

| **Primer name** | **Sequence (5′→3′)** | **Orientation** |
| --- | --- | --- |
| 8F | AGAGTTTGATCCTGGCTCAG | Forward |
| 337F | GACTCCTACGGGAGGCWGCAG | Forward |
| 518R | GTATTACCGCGGCTGCTGG | Reverse |
| 928F | TAAAACTYAAAKGAATTGACGGG | Forward |
| 1492R | CGGTTACCTTGTTACGACTT | Reverse |

#### B. PCR reaction mixture

| **Component** | **Volume (**μL**)** |
| --- | --- |
| Template DNA | 2 |
| Premix Taq Hot Start Version | 25 |
| Forward primer | 2 |
| Reverse primer | 2 |
| Distilled water | 19 |
| **Total** | **50** |

**Note**: Premix Taq Hot Start Version (Takara Bio, Shiga, Japan). Primers were synthesized by FASMAC (Atsugi, Japan). Thermal cycler: GeneAtlas Type G (Astec, Japan). Sanger sequencing was outsourced to FASMAC.

### Appendix Table 2. Tools used for whole-genome analysis

| **Tool** | **Purpose** | **Note** |
| --- | --- | --- |
| CheckM v1.2.3 | Genome quality checks | Genomes with completeness <90% or contamination ≥5% were excluded from downstream analyses*. |
| fastANI v1.34 | Average nucleotide identity (ANI) | ANI ≥95% was interpreted as the same species |
| Genome–Genome Distance Calculator 3.0 | Digital DNA-DNA hybridization (dDDH) | We used the recommended formula. Species-level relatedness was assigned at dDDH ≥70%. |
| AMRFinderPlus v4.0.23 | Antimicrobial resistance determinants | Standard thresholds were applied to classes other than β-lactamases, with the homology threshold changed to ≥30% for β-lactamases to capture distantly related homologs. |

**Note**: * Because CheckM reported 7.7% contamination even for the *D. desulfuricans* DSM 642ᵀ genome (GCA_000420465.1), we accepted contamination ≤10% for species assignment and ANI analyses within the *D. desulfuricans* lineage.

### Appendix Table 3. Detailed clinical variables and comorbidity data

| **Characteristics** | **No. (n = 8)** |
| --- | --- |
| Comorbid conditions |  |
| Myocardial infarction | 0 |
| Congestive heart failure | 3 (37.5) |
| Peripheral vascular disease | 1 (12.5) |
| Cerebrovascular disease | 1 (12.5) |
| Dementia | 0 |
| Chronic pulmonary disease | 1 (12.5) |
| Connective tissue disease | 0 |
| Peptic ulcer disease | 2 (25.0) |
| Mild liver disease | 1 (12.5) |
| Moderate or severe liver disease | 0 |
| Uncomplicated diabetes mellitus | 0 |
| Diabetes mellitus with end-organ damage | 0 |
| Hemiplegia or hemiparesis | 0 |
| Renal disease | 0 |
| Non-metastatic solid tumor | 3 (37.5) |
| Metastatic solid tumor | 0 |
| Leukemia | 0 |
| Malignant lymphoma | 1 (12.5) |
| Immunocompromised conditions |  |
| HIV | 0 |
| Chemotherapy | 1 (12.5) |
| Long-term corticosteroid use | 1 (12.5) |
| Immunosuppressive therapy | 0 |
| Organ or stem cell transplantation | 0 |
| Febrile neutropenia within 7 days | 0 |
| Indwelling devices within 30 days | 4 (50.0) |
| Symptoms |  |
| Fever | 4 (50.0) |
| Abdominal pain | 4 (50.0) |
| Abdominal distension | 1 (12.5) |
| Obstipation | 2 (25.0) |

**Footnote**: Data are no. (%) unless otherwise indicated.

### Appendix Table 4. Genome statistics of *Desulfovibrio* isolates

| **Isolate ID** | **Genome**  **size (Mb)** | **Contigs (n)** | **Longest contig (bp)** | **N50 (bp)** | **GC**  **content (%)** | **CDS**  **(n)** | **Coding**  **ratio (%)** | **rRNA (n)** | **tRNA (n)** | **CRISPR (n)** | **Completeness (%)** | **Contamination (%)** |
| --- | --- | --- | --- | --- | --- | --- | --- | --- | --- | --- | --- | --- |
| KML2501 | 2.76 | 30 | 574,094 | 265,214 | 62.4 | 2,331 | 85.7 | 3 | 47 | 2 | 99.11 | 0.01 |
| KML2502 | 3.43 | 45 | 447,596 | 215,787 | 61.4 | 2,876 | 84.8 | 3 | 51 | 1 | 99.11 | 0.00 |
| KML2503 | 3.24 | 51 | 476,950 | 138,177 | 57.7 | 2,806 | 84.1 | 3 | 48 | 1 | 98.96 | 0.00 |
| KML2504 | 3.75 | 77 | 353,551 | 164,050 | 60.8 | 3,275 | 84.4 | 3 | 48 | 0 | 98.96 | 0.22 |
| KML2505 | 3.00 | 31 | 518,440 | 214,732 | 58.0 | 2,532 | 84.1 | 3 | 48 | 1 | 98.96 | 0.00 |
| KML2506 | 3.54 | 15 | 868,136 | 358,750 | 57.2 | 3,060 | 86.1 | 3 | 54 | 1 | 93.68 | 9.60 |
| KML2507 | NA | NA | NA | NA | NA | NA | NA | NA | NA | NA | 20.61 | 0.00 |
| KML2508 | 3.54 | 94 | 300,804 | 148,734 | 57.4 | 3,101 | 85.1 | 3 | 53 | 1 | 92.71 | 10.43 |

**Footnotes**: Genome size is the total length of assembled contigs. Contig statistics and annotation were generated via the BV-BRC assembly pipeline. KML2507 did not meet inclusion criteria and is listed as NA except for the reported completeness and contamination. CDS, coding sequences; NA, not available.

### Appendix Table 5. Sequence-based analysis results

#### A. 16S rRNA BLAST results (restricted to type material)

| **Isolate ID** | **Closest match (accession)** | **Percent identity (%)** | **Identities (n/N)** | **Query**  **coverage (%)** | **E-value** |
| --- | --- | --- | --- | --- | --- |
| KML2501 | Desulfovibrio legallii H1^T^  (NR_108301.1) | 99.5 | 1,435/1,443 | 99 | 0.0 |
| KML2502 | *Desulfovibrio porci* PG-178-WT-4^T^  (MN537481.1) | 98.99 | 1,466/1,481 | 100 | 0.0 |
| KML2503 | *Desulfovibrio falkowii* 13CB8C^T^  (NR_199409.1) | 99.8 | 1,478/1,481 | 100 | 0.0 |
| KML2504 | *Desulfovibrio porci* PG-178-WT-4^T^  (MN537481.1) | 98.99 | 1,468/1,483 | 100 | 0.0 |
| KML2505 | Desulfovibrio falkowii 13CB8C^T^  (NR_199409.1) | 99.9 | 1,481/1,483 | 100 | 0.0 |
| KML2506 | *Desulfovibrio desulfuricans* Essex 6  (NR_104990.1) | 99.7 | 1,473/1,478 | 100 | 0.0 |
| KML2507 | *Desulfovibrio legallii* H1^T^  (NR_108301.1) | 99.5 | 1,435/1,443 | 97 | 0.0 |
| KML2508 | Desulfovibrio desulfuricans Essex 6  (NR_104990.1) | 99.5 | 1,471/1,478 | 100 | 0.0 |
| FH 26001/95 | Desulfovibrio porci PG-178-WT-4^T^  (MN537481.1) | 99.0 | 1,466/1,481 | 98 | 0.0 |
| D4 | Desulfovibrio porci PG-178-WT-4^T^  (MN537481.1) | 99.0 | 1,475/1,490 | 97 | 0.0 |

#### B. Sequence data used for phylogenomic analyses

| **Type strain number** | **Strain** | **Data type** | **Accession** |
| --- | --- | --- | --- |
| *D. desulfuricans* | ATCC 29577^T^ | 16S | AF192153.1 |
| *D. falkowii* | 13CB8C^T^ | 16S | LC776912.1 |
| *D. legallii* | H1^T^ | 16S | FJ225426.1 |
| *D. porci* | PG-178-WT-4^T^ | 16S | MN537481.1 |
| *D. fairfieldensi* | FH 26001/95 | 16S | U42221.1 |
| *D. fairfieldensi* | D4 | 16S | AF192155.1 |
| *NitratiDesulfovibrio liaohensis* | XJ01^T^ | 16S | MK260014.1 |
| *D. desulfuricans* | DSM 642^T^ | WGS | GCA_000420465.1 |
| *D. falkowii* | 13CB8C^T^ | WGS | GCA_045865545.1 |
| *D. legallii* | H1^T^ | WGS | GCA_004309735.1 |
| *D. porci* | PG-178-WT-4^T^ | WGS | GCA_009696265.1 |
| *D. fairfieldensi* | CCUG 45958^T^ | WGS | GCA_001553605.1 |

#### C. ANI (%) to reference genomes

| **Isolate** | ***D. desulfuricans***  **DSM 642ᵀ** | ***D. falkowii***  **13CB8Cᵀ** | ***D. legallii***  **H1ᵀ** | ***D. fairfieldensis***  **CCUG 45958** | ***D. porci***  **PG-178-WT-4ᵀ** |
| --- | --- | --- | --- | --- | --- |
| KML2501 | 79.2 | 79.3 | **99.4** | 80.1 | 80.2 |
| KML2502 | 79.3 | 79.7 | 80.2 | **98.8** | 87.4 |
| KML2503 | 79.2 | **99.2** | 78.9 | 79.7 | 78.8 |
| KML2504 | 79.1 | 79.8 | 80.2 | **98.7** | 87.3 |
| KML2505 | 79.2 | **99.4** | 78.7 | 78.7 | 78.8 |
| KML2506 | **95.5** | 79.1 | 79.4 | 79.2 | 79.1 |
| KML2508 | **95.6** | 79.2 | 79.4 | 79.3 | 79.2 |

**Footnote**: ANI computed with fastANI. Species assignment used ANI ≥95% as the same species.

#### D. Digital DNA–DNA hybridization (dDDH) results

| **Isolate ID** | **Reference species** | **Reference accession** | **dDDH**  **(%)** | **95%**  **CI (%)** | **Prob (dDDH ≥70%) (%)** | **G+C difference (%)** |
| --- | --- | --- | --- | --- | --- | --- |
| KML2502 | *D. fairfieldensi* | GCF_001553605.1 | 91.1 | 88.9 - 92.9 | 96.15 | 0.52 |
| KML2502 | *D. porci* | GCA_009696265.1 | 31.7 | 29.3 - 34.2 | 0.21 | 0.14 |
| KML2504 | *D. fairfieldensi* | GCF_001553605.1 | 90.0 | 87.7 - 91.9 | 95.81 | 0.08 |
| KML2504 | *D. porci* | GCA_009696265.1 | 31.6 | 29.2 - 34.2 | 0.2 | 0.74 |

**Footnote**: dDDH was computed with GGDC 3.0 (Formula 2; BLAST+). Values ≥70% were interpreted as the same species. Confidence intervals and probabilities are those provided by GGDC. *D. fairfieldensis* denotes a GTDB placeholder species lacking valid publication and a type strain; *D. porci* is the validly published type comparison.
